# Machine learning dimensionality reduction for heart rate n-variability (HRnV) based risk stratification of chest pain patients in the emergency department

**DOI:** 10.1101/2020.07.05.20146571

**Authors:** Nan Liu, Marcel Lucas Chee, Zhi Xiong Koh, Su Li Leow, Andrew Fu Wah Ho, Dagang Guo, Marcus Eng Hock Ong

## Abstract

**Background:** Chest pain is among the most common presenting complaints in the emergency department (ED). Swift and accurate risk stratification of chest pain patients in the ED may improve patient outcomes and reduce unnecessary costs. Traditional logistic regression with stepwise variable selection has been used to build risk prediction models for ED chest pain patients. In this study, we aimed to investigate if machine learning dimensionality reduction methods can achieve superior performance than the stepwise approach in deriving risk stratification models.

**Methods:** A retrospective analysis was conducted on the data of patients >20 years old who presented to the ED of Singapore General Hospital with chest pain between September 2010 and July 2015. Variables used included demographics, medical history, laboratory findings, heart rate variability (HRV), and HRnV parameters calculated from five to six-minute electrocardiograms (ECGs). The primary outcome was 30-day major adverse cardiac events (MACE), which included death, acute myocardial infarction, and revascularization. Candidate variables identified using univariable analysis were then used to generate the stepwise logistic regression model and eight machine learning dimensionality reduction prediction models. A separate set of models was derived by excluding troponin. Receiver operating characteristic (ROC) and calibration analysis was used to compare model performance.

**Results:** 795 patients were included in the analysis, of which 247 (31%) met the primary outcome of 30-day MACE. Patients with MACE were older and more likely to be male. All eight dimensionality reduction methods marginally but non-significantly outperformed stepwise variable selection; The multidimensional scaling algorithm performed the best with an area under the curve (AUC) of 0.901. All HRnV-based models generated in this study outperformed several existing clinical scores in ROC analysis.

**Conclusions:** HRnV-based models using stepwise logistic regression performed better than existing chest pain scores for predicting MACE, with only marginal improvements using machine learning dimensionality reduction. Moreover, traditional stepwise approach benefits from model transparency and interpretability; in comparison, machine learning dimensionality reduction models are black boxes, making them difficult to explain in clinical practice.

## Background

Chest pain is among the most common chief complaints presenting to the emergency department (ED) [1-3]. The assessment of chest pain patients poses a diagnostic challenge in balancing risk and cost. Inadvertent discharge of ACS patients is associated with higher mortality rates while inappropriate admission of patients with more benign conditions increases health service costs [4, 5]. Hence, the challenge lies in recognizing low-risk chest pain patients for safe and early discharge from the ED. There has been increasing focus on the development of risk stratification scores. Initially, risk scores such as the Thrombolysis in Myocardial Infarction (TIMI) score [6, 7] and the Global Registry of Acute Coronary Events (GRACE) score [8] were developed from post-ACS patients to estimate short-term mortality and recurrence of myocardial infarction. The History, Electrocardiogram (ECG), Age, Risk factors, and initial Troponin (HEART) score was subsequently designed for ED chest pain patients [9], which demonstrated superior performance in many comparison studies on the identification of low-risk chest pain patients [10-17]. Nonetheless, the HEART score has its disadvantages. Many potential factors can affect its diagnostic and prognostic accuracy, such as variation in patient populations, provider determination of low-risk heart score criteria, specific troponin reagent used, all of which contribute to clinical heterogeneity [18-21]. In addition, most risk scores still require variables that may not be available during the initial presentation of the patient to the ED such as troponin. There remains a need for a more efficient risk stratification tool.

We had previously developed a heart rate variability (HRV) prediction model using readily available variables at the ED, in an attempt to reduce both diagnostic time and subjective components [22]. HRV characterizes beat-to-beat variation using time, frequency domain and nonlinear analysis [23], and has proven to be a good predictor of major adverse cardiac events (MACE) [22, 24, 25]. Most HRV-based scores were reported to be superior to TIMI and GRACE scores while achieving comparable performance with HEART score [17, 24, 26, 27]. Recently, we established a new representation of beat-to-beat variation in ECGs, the heart rate n-variability (HRnV) [28]. HRnV utilises variation in sampling RR-intervals and overlapping RR-intervals to derive additional parameters from a single strip of ECG reading. As an extension to HRV, HRnV potentially supplements additional information about adverse cardiac events while reducing unwanted noise caused by abnormal heart beats. Moreover, HRV is a special case of HRnV when *n* = 1. The HRnV prediction model, developed from multivariable stepwise logistic regression, outperformed the HEART, TIMI and GRACE scores in predicting 30-day MACE [28]. Nevertheless, multicollinearity is a common problem in logistic regression models where supposedly independent predictor variables are correlated. They tend to overestimate the variance of regression parameters and hinder the determination of the exact effect of each parameter, which could potentially result in inaccurate identification of significant predictors [29, 30]. In the paper, 115 HRnV parameters were derived but only seven variables were left in the final prediction model, and this implies the possible elimination of relevant information [28].

Within the general medical literature, machine learning dimensionality reduction methods are uncommon and limited to a few specific areas, such as bioinformatics studies on genetics [31, 32] and diagnostic radiological imaging [33, 34]. Despite this, dimensionality reduction in HRV has been investigated and shown to effectively compress multidimensional HRV data for assessment of cardiac autonomic neuropathy [35]. In this paper, we attempted to investigate several machine learning dimensionality reduction algorithms as possible alternatives to stepwise selection, hypothesizing that these algorithms could be useful in preserving useful information while improving prediction performance. We aimed to compare the performance of HRnV-based dimensionality reduction models against HRnV stepwise logistic regression model and conventional risk stratification tools such as the HEART, TIMI, and GRACE scores, in the prediction of 30-day MACE in chest pain patients presenting to the ED.

## Methods

### Study design and clinical setting

A retrospective analysis was conducted on data collected from patients >20 years old who presented to Singapore General Hospital ED with chest pain between September 2010 to July 2015. These patients were triaged using the Patient Acuity Category Scale (PACS) and those with PACS 1 or 2 were included in the study. Patients were excluded if they were lost to the 30-day follow-up or if they presented with ST-elevation myocardial infarction (STEMI) or non-cardiac aetiology chest pain such as pneumothorax, pneumonia and trauma as diagnosed by the ED physician. Patients with ECG findings that precluded quality HRnV analysis such as artefacts, ectopic beats, paced or non-sinus rhythm were also excluded.

### Data collection

For each patient, HRV and HRnV parameters were calculated using HRnV-Calc software suite [28, 36] from a five to six-minute single-lead (lead II) ECG performed via the X-series Monitor (ZOLL Medical, Corporation, Chelmsford, MA). Table 1 shows the full list of HRV and HRnV parameters used in this study. In addition, the first 12-lead ECGs taken during patients’ presentation to the ED was interpreted by two independent clinical reviewers and any pathological ST changes, T wave inversions and Q-waves were noted. Patients’ demographics, medical history, first set of vital signs and troponin-T values were obtained from the hospital’s electronic health records (EHR). In this study, high-sensitivity troponin-T was selected as the cardiac biomarker and an abnormal value was defined as >0.03ng/mL.

**Table 1:** List of traditional heart rate variability (HRV) and novel heart rate n-variability (HRnV) parameters used in this study.

| HRV | HR<sub>2</sub>V | HR<sub>2</sub>V<sub>1</sub> | HR<sub>3</sub>V | HR<sub>3</sub>V<sub>1</sub> | HR<sub>3</sub>V<sub>2</sub> |
| --- | --- | --- | --- | --- | --- |
| Mean NN | HR<sub>2</sub>V Mean NN | HR<sub>2</sub>V<sub>1</sub> Mean NN | HR<sub>3</sub>V Mean NN | HR<sub>3</sub>V<sub>1</sub> Mean NN | HR<sub>3</sub>V<sub>2</sub> Mean NN |
| SDNN | HR<sub>2</sub>V SDNN | HR<sub>2</sub>V<sub>1</sub> SDNN | HR<sub>3</sub>V SDNN | HR<sub>3</sub>V<sub>1</sub> SDNN | HR<sub>3</sub>V<sub>2</sub> SDNN |
| RMSSD | HR<sub>2</sub>V RMSSD | HR<sub>2</sub>V<sub>1</sub> RMSSD | HR<sub>3</sub>V RMSSD | HR<sub>3</sub>V<sub>1</sub> RMSSD | HR<sub>3</sub>V<sub>2</sub> RMSSD |
| Skewness | HR<sub>2</sub>V Skewness | HR<sub>2</sub>V<sub>1</sub> Skewness | HR<sub>3</sub>V Skewness | HR<sub>3</sub>V<sub>1</sub> Skewness | HR<sub>3</sub>V<sub>2</sub> Skewness |
| Kurtosis | HR<sub>2</sub>V Kurtosis | HR<sub>2</sub>V<sub>1</sub> Kurtosis | HR<sub>3</sub>V Kurtosis | HR<sub>3</sub>V<sub>1</sub> Kurtosis | HR<sub>3</sub>V<sub>2</sub> Kurtosis |
| Triangular index | HR<sub>2</sub>V Triangular index | HR<sub>2</sub>V<sub>1</sub> Triangular index | HR<sub>3</sub>V Triangular index | HR<sub>3</sub>V<sub>1</sub> Triangular index | HR<sub>3</sub>V<sub>2</sub> Triangular index |
| NN50 | HR<sub>2</sub>V NN50 | HR<sub>2</sub>V<sub>1</sub> NN50 | HR<sub>3</sub>V NN50 | HR<sub>3</sub>V<sub>1</sub> NN50 | HR<sub>3</sub>V<sub>2</sub> NN50 |
| pNN50 | HR<sub>2</sub>V pNN50 | HR<sub>2</sub>V<sub>1</sub> pNN50 | HR<sub>3</sub>V pNN50 | HR<sub>3</sub>V<sub>1</sub> pNN50 | HR<sub>3</sub>V<sub>2</sub> pNN50 |
| - | HR<sub>2</sub>V NN50<sub>n</sub> | HR<sub>2</sub>V<sub>1</sub> NN50<sub>n</sub> | HR<sub>3</sub>V NN50<sub>n</sub> | HR<sub>3</sub>V<sub>1</sub> NN50<sub>n</sub> | HR<sub>3</sub>V<sub>2</sub> NN50<sub>n</sub> |
| - | HR<sub>2</sub>V pNN50<sub>n</sub> | HR<sub>2</sub>V<sub>1</sub> pNN50<sub>n</sub> | HR<sub>3</sub>V pNN50<sub>n</sub> | HR<sub>3</sub>V<sub>1</sub> pNN50<sub>n</sub> | HR<sub>3</sub>V<sub>2</sub> pNN50<sub>n</sub> |
| Total power | HR<sub>2</sub>V Total power | HR<sub>2</sub>V<sub>1</sub> Total power | HR<sub>3</sub>V Total power | HR<sub>3</sub>V<sub>1</sub> Total power | HR<sub>3</sub>V<sub>2</sub> Total power |
| VLF power | HR<sub>2</sub>V VLF power | HR<sub>2</sub>V<sub>1</sub> VLF power | HR<sub>3</sub>V VLF power | HR<sub>3</sub>V<sub>1</sub> VLF power | HR<sub>3</sub>V<sub>2</sub> VLF power |
| LF power | HR<sub>2</sub>V LF power | HR<sub>2</sub>V<sub>1</sub> LF power | HR<sub>3</sub>V LF power | HR<sub>3</sub>V<sub>1</sub> LF power | HR<sub>3</sub>V<sub>2</sub> LF power |
| HF power | HR<sub>2</sub>V HF power | HR<sub>2</sub>V<sub>1</sub> HF power | HR<sub>3</sub>V HF power | HR<sub>3</sub>V<sub>1</sub> HF power | HR<sub>3</sub>V<sub>2</sub> HF power |
| LF power norm | HR<sub>2</sub>V LF power norm | HR<sub>2</sub>V<sub>1</sub> LF power norm | HR<sub>3</sub>V LF power norm | HR<sub>3</sub>V<sub>1</sub> LF power norm | HR<sub>3</sub>V<sub>2</sub> LF power norm |
| HF power norm | HR<sub>2</sub>V HF power norm | HR<sub>2</sub>V<sub>1</sub> HF power norm | HR<sub>3</sub>V HF power norm | HR<sub>3</sub>V<sub>1</sub> HF power norm | HR<sub>3</sub>V<sub>2</sub> HF power norm |
| LF/HF | HR<sub>2</sub>V LF/HF | HR<sub>2</sub>V<sub>1</sub> LF/HF | HR<sub>3</sub>V LF/HF | HR<sub>3</sub>V<sub>1</sub> LF/HF | HR<sub>3</sub>V<sub>2</sub> LF/HF |
| Poincaré SD1 | HR<sub>2</sub>V Poincaré SD1 | HR<sub>2</sub>V<sub>1</sub> Poincaré SD1 | HR<sub>3</sub>V Poincaré SD1 | HR<sub>3</sub>V<sub>1</sub> Poincaré SD1 | HR<sub>3</sub>V<sub>2</sub> Poincaré SD1 |
| Poincaré SD2 | HR<sub>2</sub>V Poincaré SD2 | HR<sub>2</sub>V<sub>1</sub> Poincaré SD2 | HR<sub>3</sub>V Poincaré SD2 | HR<sub>3</sub>V<sub>1</sub> Poincaré SD2 | HR<sub>3</sub>V<sub>2</sub> Poincaré SD2 |
| Poincaré SD1/SD2 ratio | HR<sub>2</sub>V Poincaré SD1/SD2 | HR<sub>2</sub>V<sub>1</sub> Poincaré SD1/SD2 | HR<sub>3</sub>V Poincaré SD1/SD2 | HR<sub>3</sub>V<sub>1</sub> Poincaré SD1/SD2 | HR<sub>3</sub>V<sub>2</sub> Poincaré SD1/SD2 |
| SampEn | HR<sub>2</sub>V SampEn | HR<sub>2</sub>V<sub>1</sub> SampEn | HR<sub>3</sub>V SampEn | HR<sub>3</sub>V<sub>1</sub> SampEn | HR<sub>3</sub>V<sub>2</sub> SampEn |
| ApEn | HR<sub>2</sub>V ApEn | HR<sub>2</sub>V<sub>1</sub> ApEn | HR<sub>3</sub>V ApEn | HR<sub>3</sub>V<sub>1</sub> ApEn | HR<sub>3</sub>V<sub>2</sub> ApEn |
| DFA, $\alpha_1$ | HR<sub>2</sub>V DFA, $\alpha_1$ | HR<sub>2</sub>V<sub>1</sub> DFA, $\alpha_1$ | HR<sub>3</sub>V DFA, $\alpha_1$ | HR<sub>3</sub>V<sub>1</sub> DFA, $\alpha_1$ | HR<sub>3</sub>V<sub>2</sub> DFA, $\alpha_1$ |
| DFA, $\alpha_2$ | HR<sub>2</sub>V DFA, $\alpha_2$ | HR<sub>2</sub>V<sub>1</sub> DFA, $\alpha_2$ | HR<sub>3</sub>V DFA, $\alpha_2$ | HR<sub>3</sub>V<sub>1</sub> DFA, $\alpha_2$ | HR<sub>3</sub>V<sub>2</sub> DFA, $\alpha_2$ |

Mean NN, average of R-R intervals; SDNN, standard deviation of R-R intervals; RMSSD, square root of the mean squared differences between R-R intervals; NN50, the number of times that the absolute difference between 2 successive R-R intervals exceeds 50 ms; pNN50, NN50 divided by the total number of R-R intervals; NN50<sub>n</sub>, the number of times that the absolute difference between 2 successive RR<sub>n</sub>/RR<sub>n</sub>I<sub>m</sub> sequences exceeds 50×*n* ms; pNN50<sub>n</sub>, NN50<sub>n</sub> divided by the total number of RR<sub>n</sub>/RR<sub>n</sub>I<sub>m</sub> sequences; VLF, very low frequency; LF, low frequency; HF, high frequency; SD, standard deviation; SampEn, sample entropy; ApEn, approximate entropy; DFA, detrended fluctuation analysis.

The primary outcome measured was any MACE within 30 days, including acute myocardial infarction (STEMI/Non-STEMI), emergent revascularization procedures such as percutaneous coronary intervention (PCI) or coronary artery bypass graft (CABG), or death. The primary outcome was captured through retrospective review of patients’ EHR.

### Machine learning dimensionality reduction

Dimensionality reduction in machine leaning and data mining [37] refers to the process of transforming high-dimensional data into lower dimensions such that less features are selected or extracted while preserving essential information of the original data. Two types of dimensionality reduction approaches are available, namely feature selection (commonly called variable selection in biostatistics) and feature extraction. Feature selection methods generally reduce data dimensionality by choosing a subset of variables, while feature extraction methods transform the original feature space into lower-dimensional space through linear or nonlinear feature projection. In clinical predictive modelling, feature selection techniques such as stepwise logistic regression are popular for constructing prediction models [38]. In contrast, feature extraction approaches [39] are less commonly used in medical research, although they have been widely used in computational biology [40], image analysis [41, 42], physiological signal analysis [43], among others. In this study, we investigated the implementation of eight feature extraction algorithms and evaluated their contributions to prediction performance in risk stratification of ED chest pain patients. We also compared them with a prediction model that was built using conventional stepwise variable selection [28]. Henceforth, we use the terms “dimensionality reduction” and “feature extraction” interchangeably.

Given that there were *n* samples (*x*_*i*_, *y*_*i*_), *i* = 1,2, …, *n*, in the dataset (***X, y***), where each sample *x*_*i*_ had original *D* features/variables and its label *y*_*i*_ = 1 or 0, with 1 indicating a positive primary outcome, i.e. MACE within 30 days. We applied dimensionality reduction algorithms to project *x*_*i*_ into a *d*-dimensional space (*d* < *D*). As a result, the original dataset ***X*** ∈ ℝ^*n*×*D*^ became 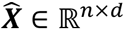. There was a total of *D* = 174 candidate variables in this study. As suggested in Liu et al. [28], some variables were less statistically significant in terms of contributions to the prediction performance. Thus, we conducted univariable analysis and preselected a subset of 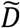 variables if their 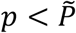, where 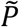 was a threshold determined by predictive performance using PCA [44], the most common technique for linear dimensionality reduction. In this study, we implemented eight dimensionality reduction algorithms, including PCA, kernel PCA (KPCA) [45] with polynomial kernel function, latent semantic analysis (LSA) [46], Gaussian random projection (GRP) [47], sparse random projection (SRP) [48], multidimensional scaling (MDS) [49], Isomap [50], and locally linear embedding (LLE) [51]. All these algorithms are unsupervised learning methods, meaning the transformation of feature space does not rely on sample labels ***y***. Among the eight methods, MDS, Isomap, and LLE are manifold learning-based techniques for nonlinear dimensionality reduction.

### Predictive and statistical analysis

In this study, we chose logistic regression as the classification algorithm to predict the MACE outcome. Before applying all eight dimensionality reduction algorithms, we ran PCA and logistic regression through 5-fold cross-validation to determine the threshold 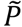 to preselect a subset of 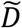 variables, such that the prediction performance in terms of area under the curve (AUC) was the largest. We did this preselection to ensure the removal of less significant variables as shown by univariable analysis, after which ***X*** ∈ ℝ^*n*×*D*^ became 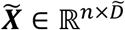. In summary, the inputs to all dimensionality reduction algorithms were in 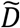-dimensional space. Subsequently, conventional logistic regression was implemented to take *d*-dimensional 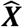 to predict ***y***, where 5-fold cross-validation was used.

To compare machine learning dimensionality reduction and traditional stepwise variable selection, we implemented our previously built logistic regression model [28], in which the following 16 variables were used: age, diastolic blood pressure, pain score, ST-elevation, ST-depression, Q wave, cardiac history (the “History” component in the HEART score), troponin, HRV NN50, HR_2_V skewness, HR_2_V SampEn, HR_2_V ApEn, HR_2_V_1_ ApEn, HR_3_V RMSSD, HR_3_V skewness, and HR_3_V_2_ HF power. The predictive performances of eight models built from different dimensionality reduction algorithms and the model obtained using convention stepwise variable selection [28] were compared with the receiver operating characteristic (ROC) curve analysis, and the corresponding sensitivity, specificity, positive predictive value (PPV), and negative predictive value (NPV) measures were reported. The calibration plots of all nine prediction models were also generated. All analyses were conducted in Python version 3.8.0 (Python Software Foundation, Delaware, USA).

## Results

We included 795 chest pain patients in this study, of which 247 (31%) patients had MACE within 30 days of presentation to the ED. Table 1 presents the baseline characteristics of the patient cohort. Patients with MACE were older (median age 61 years vs. 59 years, p = 0.035) and more likely to be male (76.1% vs. 64.6%, p = 0.002). History of diabetes, current smoking status, and pathological ECG changes such as ST elevation, ST depression, T wave inversion, pathological Q waves, and QTc prolongation were significantly more prevalent in patients with the primary outcome. Troponin-T and creatine kinase-MB levels were also significantly elevated in patients with the primary outcome. There was no statistically significant difference in patient ethnicity between MACE and non-MACE groups.

Figure 1(a) depicts the PCA-based predictive performance versus the threshold 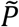 (for preselection of variables) and Figure 1(b) shows the number of preselected variables versus threshold 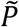. The predictive performance peaked at 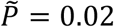, where a total of 30 variables were preselected, including gender, diastolic blood pressure, pain score, ST-elevation, ST-depression, T-wave inversion, Q wave, cardiac history, EKG and risk factor components of the HEART score, troponin, HRV RMSSD, HRV NN50, HRV pNN50, HRV HF power, HRV Poincaré SD1, HR_2_V RMSSD, HR_2_V NN50, HR_2_V pNN50, HR_2_V HF power, HR_2_V Poincaré SD1, HR_2_V_1_ RMSSD, HR_2_V_1_ NN50, HR_2_V_1_ HF power, HR_2_V_1_ Poincaré SD1, HR_3_V_1_ RMSSD, HR_3_V_1_ HF power, HR_3_V_1_ Poincaré SD1, HR_3_V_2_ RMSSD, and HR_3_V_2_ Poincaré SD1. These were used as inputs to all dimensionality reduction algorithms whose outputs were linear or nonlinear combinations of these 30 variables.

**Figure 1:**
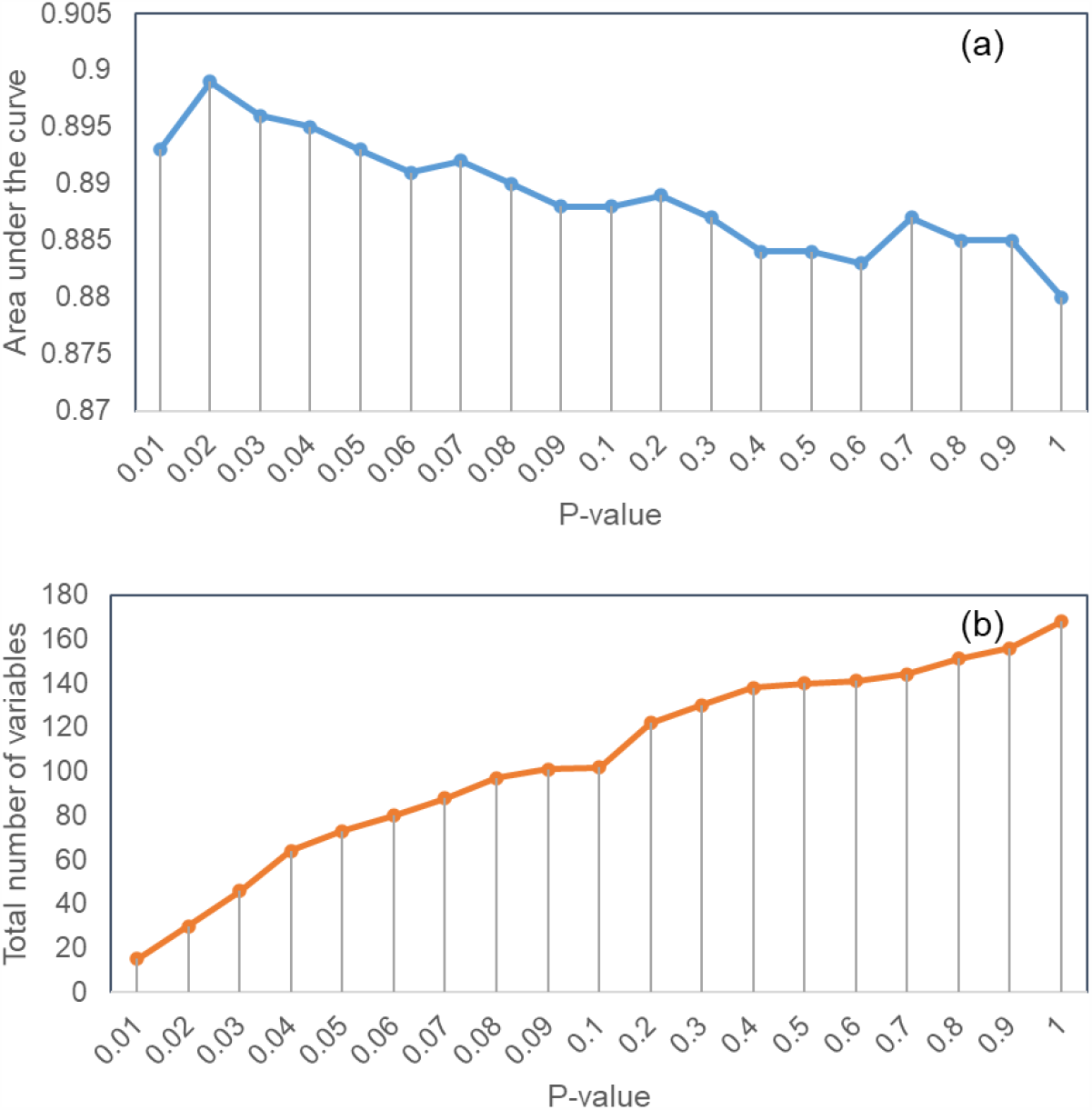
Variable preselection using p-value in univariable analysis for dimensionality reduction: (a) prediction area under the curve versus the p-value, and (b) the number of preselected variables versus the p-value.

Figure 2 shows the predictive performance (in terms of AUC value) versus feature dimension (i.e. number of “principal components”) for all eight dimensionality reduction algorithms. The AUC values of GRP, SRP, and KPCA gradually increased with the increment of feature dimension, while the AUC values of PCA, LSA, MDS, Isomap, and LLE drastically jumped to more than 0.8 when feature dimension *d* ≥ 3 and plateaued in the curves when *d* ≥ 15. The highest AUC values of PCA, KPCA, LSA, GRP, SRP, MDS, Isomap, and LLE were 0.899, 0.896, 0.899, 0.896, 0.898, 0.901, 0.888, and 0.898, achieved with feature dimensions of 15, 30, 15, 22, 20, 27, 23, and 30, respectively.

**Figure 2:**
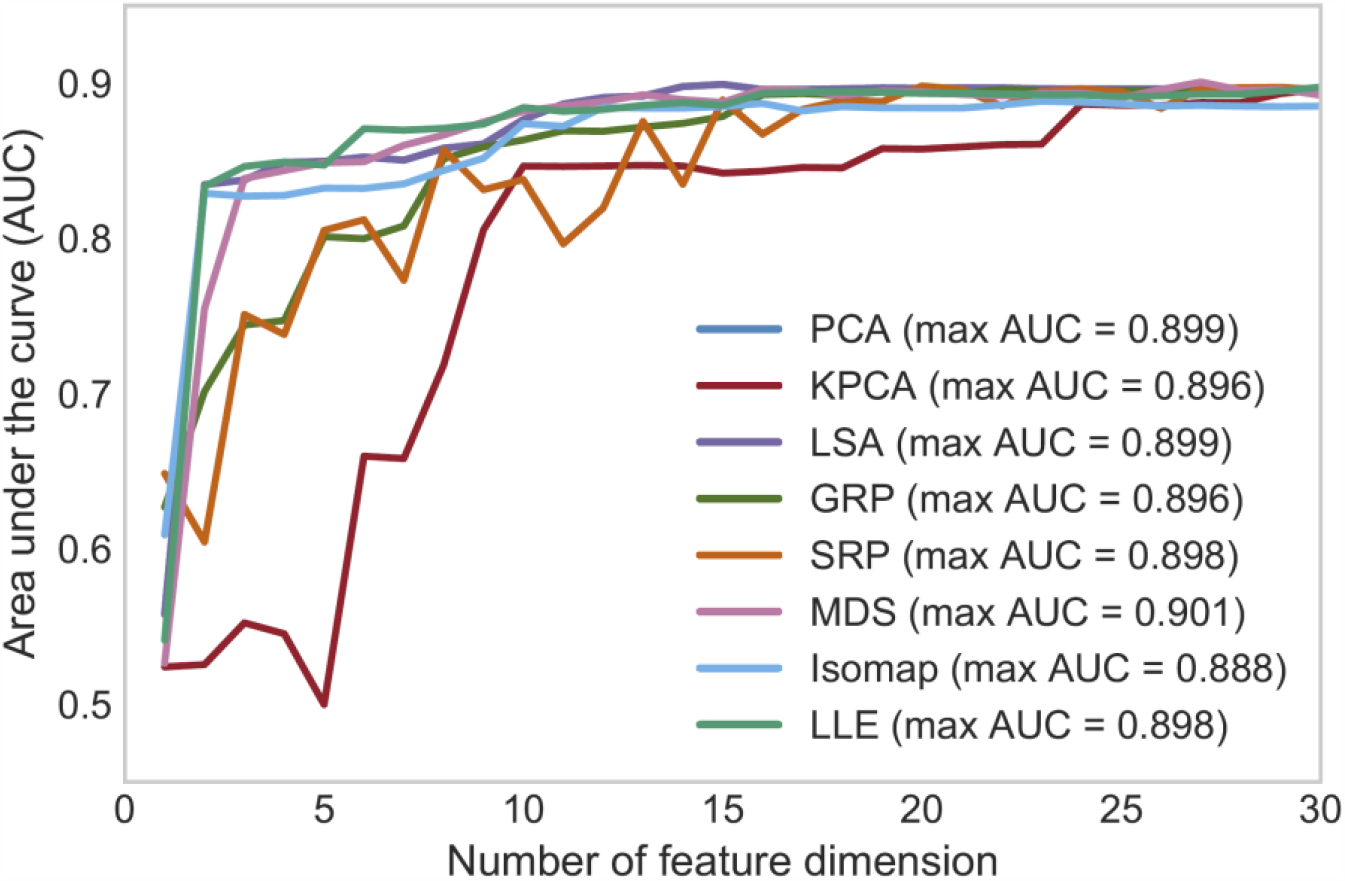
Prediction performance based on the eight dimensionality reduction algorithms versus the number of feature dimensions after reduction.

Figure 3 shows the ROC curves of the eight dimensionality reduction algorithms, the stepwise logistic regression, and three clinical scores. All eight dimensionality reduction methods marginally outperformed the stepwise variable selection, and MDS performed the best with an AUC of 0.901. Table 3 presents ROC analysis results of all 12 methods/scores where sensitivity, specificity, PPV, and NPV are reported with 95% confidence intervals (CIs). Figure 4 presents the calibration curves of predictions by all methods/scores. The stepwise model and seven dimensionality reduction models (PCA, KPCA, LSA, GRP, SRP, MDS, and Isomap) showed reasonable model calibrations, in which their curves fluctuated along the diagonal line, meaning these models only slightly overestimated or underestimated the predicted probability of 30-day MACE. The LLE model was unable to achieve good calibration. In comparison, all three clinical scores (HEART, TIMI, and GRACE) generally underpredicted the probability of 30-day MACE.

**Table 2:**
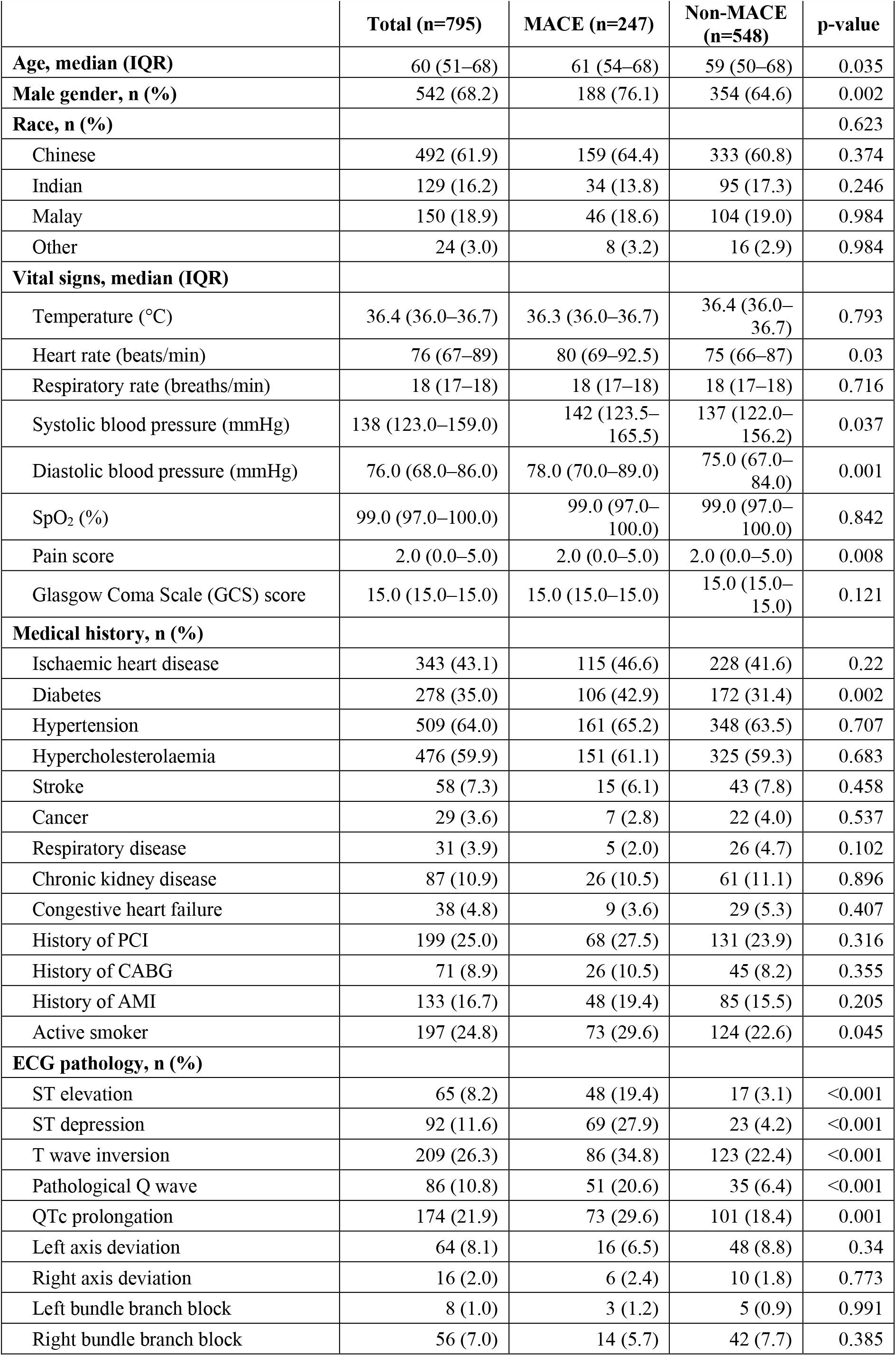
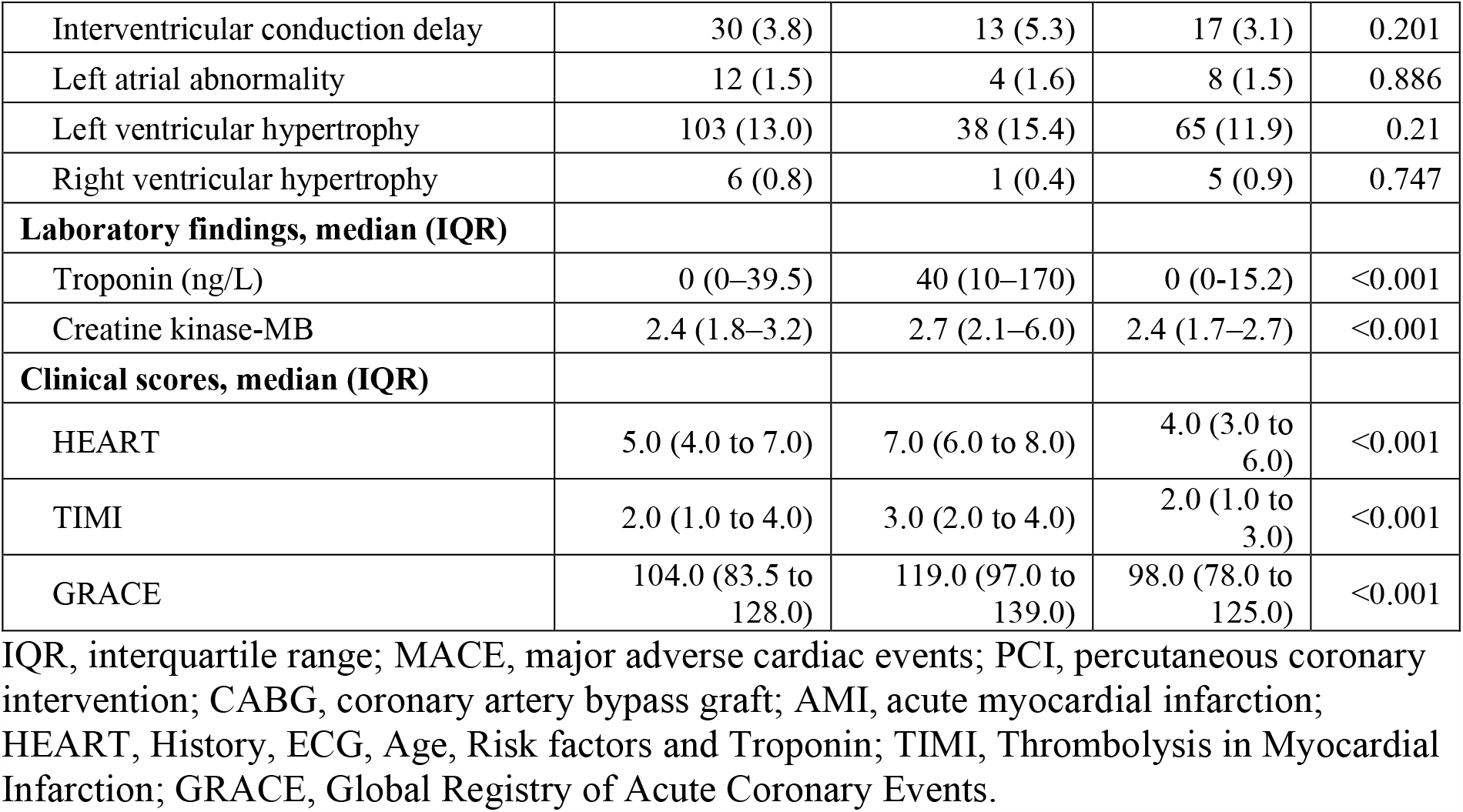
Baseline characteristics of patient cohorts.

|  | **Total (n=795)** | **MACE (n=247)** | **Non-MACE (n=548)** | **p-value** |
| --- | --- | --- | --- | --- |
| **Age, median (IQR)** | 60 (51–68) | 61 (54–68) | 59 (50–68) | 0.035 |
| **Male gender, n (%)** | 542 (68.2) | 188 (76.1) | 354 (64.6) | 0.002 |
| **Race, n (%)** |  |  |  | 0.623 |
| Chinese | 492 (61.9) | 159 (64.4) | 333 (60.8) | 0.374 |
| Indian | 129 (16.2) | 34 (13.8) | 95 (17.3) | 0.246 |
| Malay | 150 (18.9) | 46 (18.6) | 104 (19.0) | 0.984 |
| Other | 24 (3.0) | 8 (3.2) | 16 (2.9) | 0.984 |
| **Vital signs, median (IQR)** |  |  |  |  |
| Temperature (°C) | 36.4 (36.0–36.7) | 36.3 (36.0–36.7) | 36.4 (36.0–36.7) | 0.793 |
| Heart rate (beats/min) | 76 (67–89) | 80 (69–92.5) | 75 (66–87) | 0.03 |
| Respiratory rate (breaths/min) | 18 (17–18) | 18 (17–18) | 18 (17–18) | 0.716 |
| Systolic blood pressure (mmHg) | 138 (123.0–159.0) | 142 (123.5–165.5) | 137 (122.0–156.2) | 0.037 |
| Diastolic blood pressure (mmHg) | 76.0 (68.0–86.0) | 78.0 (70.0–89.0) | 75.0 (67.0–84.0) | 0.001 |
| SpO<sub>2</sub> (%) | 99.0 (97.0–100.0) | 99.0 (97.0–100.0) | 99.0 (97.0–100.0) | 0.842 |
| Pain score | 2.0 (0.0–5.0) | 2.0 (0.0–5.0) | 2.0 (0.0–5.0) | 0.008 |
| Glasgow Coma Scale (GCS) score | 15.0 (15.0–15.0) | 15.0 (15.0–15.0) | 15.0 (15.0–15.0) | 0.121 |
| **Medical history, n (%)** |  |  |  |  |
| Ischaemic heart disease | 343 (43.1) | 115 (46.6) | 228 (41.6) | 0.22 |
| Diabetes | 278 (35.0) | 106 (42.9) | 172 (31.4) | 0.002 |
| Hypertension | 509 (64.0) | 161 (65.2) | 348 (63.5) | 0.707 |
| Hypercholesterolaemia | 476 (59.9) | 151 (61.1) | 325 (59.3) | 0.683 |
| Stroke | 58 (7.3) | 15 (6.1) | 43 (7.8) | 0.458 |
| Cancer | 29 (3.6) | 7 (2.8) | 22 (4.0) | 0.537 |
| Respiratory disease | 31 (3.9) | 5 (2.0) | 26 (4.7) | 0.102 |
| Chronic kidney disease | 87 (10.9) | 26 (10.5) | 61 (11.1) | 0.896 |
| Congestive heart failure | 38 (4.8) | 9 (3.6) | 29 (5.3) | 0.407 |
| History of PCI | 199 (25.0) | 68 (27.5) | 131 (23.9) | 0.316 |
| History of CABG | 71 (8.9) | 26 (10.5) | 45 (8.2) | 0.355 |
| History of AMI | 133 (16.7) | 48 (19.4) | 85 (15.5) | 0.205 |
| Active smoker | 197 (24.8) | 73 (29.6) | 124 (22.6) | 0.045 |
| **ECG pathology, n (%)** |  |  |  |  |
| ST elevation | 65 (8.2) | 48 (19.4) | 17 (3.1) | <0.001 |
| ST depression | 92 (11.6) | 69 (27.9) | 23 (4.2) | <0.001 |
| T wave inversion | 209 (26.3) | 86 (34.8) | 123 (22.4) | <0.001 |
| Pathological Q wave | 86 (10.8) | 51 (20.6) | 35 (6.4) | <0.001 |
| QTc prolongation | 174 (21.9) | 73 (29.6) | 101 (18.4) | 0.001 |
| Left axis deviation | 64 (8.1) | 16 (6.5) | 48 (8.8) | 0.34 |
| Right axis deviation | 16 (2.0) | 6 (2.4) | 10 (1.8) | 0.773 |
| Left bundle branch block | 8 (1.0) | 3 (1.2) | 5 (0.9) | 0.991 |
| Right bundle branch block | 56 (7.0) | 14 (5.7) | 42 (7.7) | 0.385 |
| Interventricular conduction delay | 30 (3.8) | 13 (5.3) | 17 (3.1) | 0.201 |
| Left atrial abnormality | 12 (1.5) | 4 (1.6) | 8 (1.5) | 0.886 |
| Left ventricular hypertrophy | 103 (13.0) | 38 (15.4) | 65 (11.9) | 0.21 |
| Right ventricular hypertrophy | 6 (0.8) | 1 (0.4) | 5 (0.9) | 0.747 |
| **Laboratory findings, median (IQR)** |  |  |  |  |
| Troponin (ng/L) | 0 (0–39.5) | 40 (10–170) | 0 (0–15.2) | <0.001 |
| Creatine kinase-MB | 2.4 (1.8–3.2) | 2.7 (2.1–6.0) | 2.4 (1.7–2.7) | <0.001 |
| **Clinical scores, median (IQR)** |  |  |  |  |
| HEART | 5.0 (4.0 to 7.0) | 7.0 (6.0 to 8.0) | 4.0 (3.0 to 6.0) | <0.001 |
| TIMI | 2.0 (1.0 to 4.0) | 3.0 (2.0 to 4.0) | 2.0 (1.0 to 3.0) | <0.001 |
| GRACE | 104.0 (83.5 to 128.0) | 119.0 (97.0 to 139.0) | 98.0 (78.0 to 125.0) | <0.001 |

IQR, interquartile range; MACE, major adverse cardiac events; PCI, percutaneous coronary intervention; CABG, coronary artery bypass graft; AMI, acute myocardial infarction; HEART, History, ECG, Age, Risk factors and Troponin; TIMI, Thrombolysis in Myocardial Infarction; GRACE, Global Registry of Acute Coronary Events.

**Table 3:** Comparison of performance of the HRnV models (based on 5-fold cross-validation), HEART, TIMI, and GRACE scores in predicting 30-day major adverse cardiac events (MACE). The cut-off values were defined as the points nearest to the upper-left corner on the ROC curves.

|  | AUC (95% CI) | Cut-off | Sensitivity (95% CI) | Specificity (95% CI) | PPV (95% CI) | NPV (95% CI) |
| --- | --- | --- | --- | --- | --- | --- |
| **Stepwise** | 0.887 (0.859-0.916) | 0.3140 | 79.4% (74.3% - 84.4%) | 78.8% (75.4% - 82.3%) | 62.8% (57.5% - 68.2%) | 89.4% (86.7% - 92.2%) |
| **PCA** | 0.899 (0.872-0.926) | 0.2881 | 85.4% (81.0% - 89.8%) | 78.5% (75.0% - 81.9%) | 64.1% (59.0% - 69.3%) | 92.3% (89.9% - 94.7%) |
| **KPCA** | 0.896 (0.869-0.923) | 0.3489 | 81.8% (77.0% - 86.6%) | 82.1% (78.9% - 85.3%) | 67.3% (62.0% - 72.6%) | 90.9% (88.4% - 93.4%) |
| **LSA** | 0.899 (0.872-0.926) | 0.2884 | 85.4% (81.0% - 89.8%) | 78.6% (75.2% - 82.1%) | 64.3% (59.1% - 69.5%) | 92.3% (89.9% - 94.7%) |
| **GRP** | 0.896 (0.868-0.923) | 0.2965 | 85.0% (80.6% - 89.5%) | 78.5% (75.0% - 81.9%) | 64.0% (58.8% - 69.2%) | 92.1% (89.6% - 94.5%) |
| **SRP** | 0.898 (0.871-0.925) | 0.2940 | 84.6% (80.1% - 89.1%) | 79.6% (76.2% - 82.9%) | 65.1% (59.9% - 70.3%) | 92.0% (89.5% - 94.4%) |
| **MDS** | 0.901 (0.874-0.928) | 0.3095 | 83.4% (78.8% - 88.0%) | 81.6% (78.3% - 84.8%) | 67.1% (61.8% - 72.4%) | 91.6% (89.1% - 94.1%) |
| **Isomap** | 0.888 (0.860-0.917) | 0.3468 | 78.5% (73.4% - 83.7%) | 82.7% (79.5% - 85.8%) | 67.1% (61.7% - 72.5%) | 89.5% (86.9% - 92.2%) |
| **LLE** | 0.898 (0.870-0.925) | 0.3140 | 85.0% (80.6% - 89.5%) | 79.4% (76.0% - 82.8%) | 65.0% (59.8% - 70.2%) | 92.2% (89.7% - 94.6%) |
| **HEART** | 0.841 (0.808-0.874) | 5 | 78.9% (73.9% - 84.0%) | 72.8% (69.1% - 76.5%) | 56.7% (51.4% - 61.9%) | 88.5% (85.5% - 91.4%) |
| **TIMI** | 0.681 (0.639-0.723) | 2 | 63.6% (57.6% - 69.6%) | 58.4% (54.3% - 62.5%) | 40.8% (35.9% - 45.7%) | 78.0% (74.0% - 82.1%) |
| **GRACE** | 0.665 (0.623-0.707) | 107 | 64.0% (58.0% - 70.0%) | 60.8% (56.7% - 64.9%) | 42.4% (37.3% - 47.4%) | 78.9% (75.0% - 82.8%) |

AUC, area under the curve; CI, confidence interval; PPV, positive predictive value; NPV, negative predictive value; HEART, History, ECG, Age, Risk factors and Troponin; TIMI, Thrombolysis in Myocardial Infarction; GRACE, Global Registry of Acute Coronary Events.

**Figure 3:**
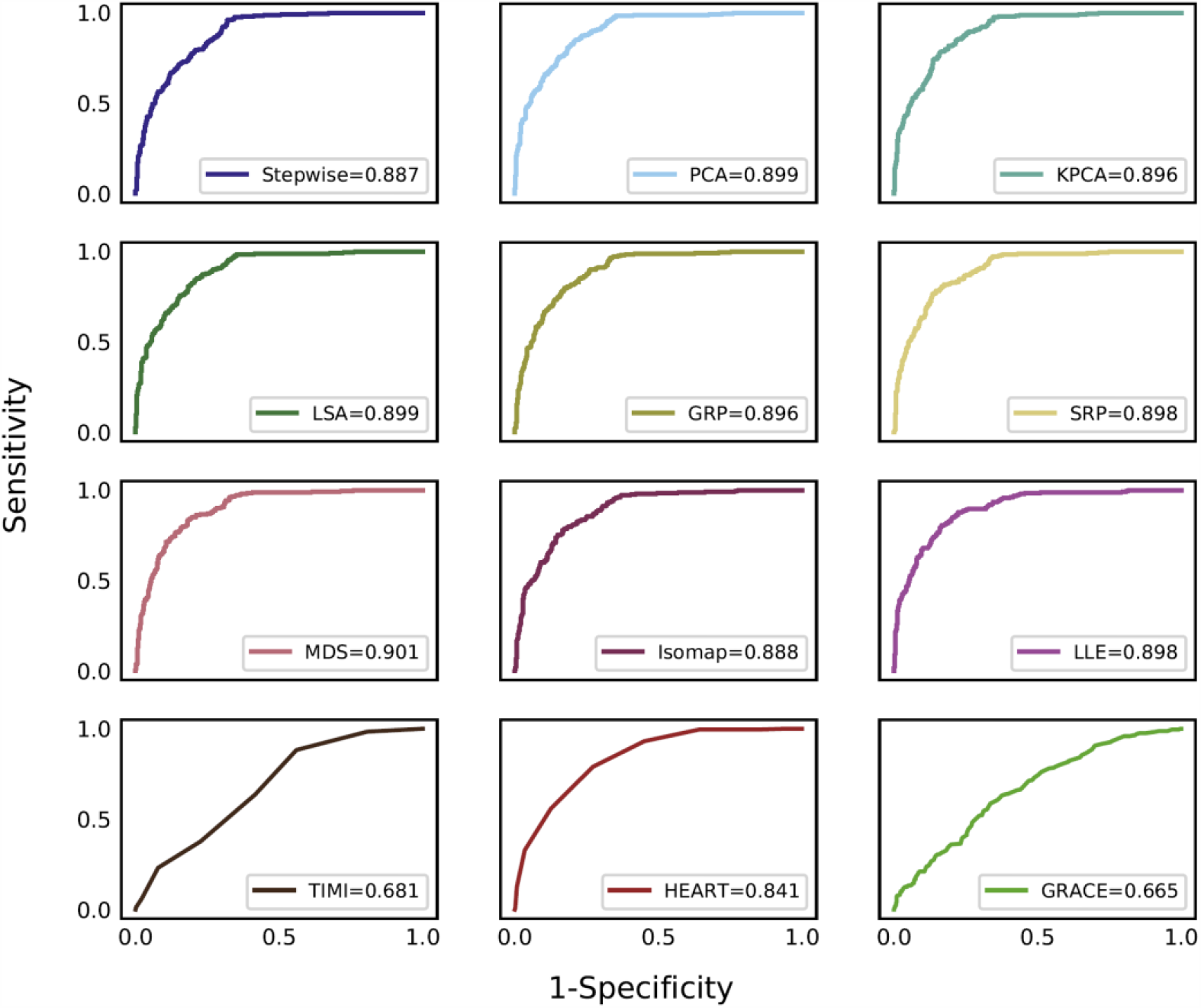
ROC curves (based on the optimal number of dimensions) generated by the stepwise model, eight dimensionality reduction models, and three clinical scores.

**Figure 4:**
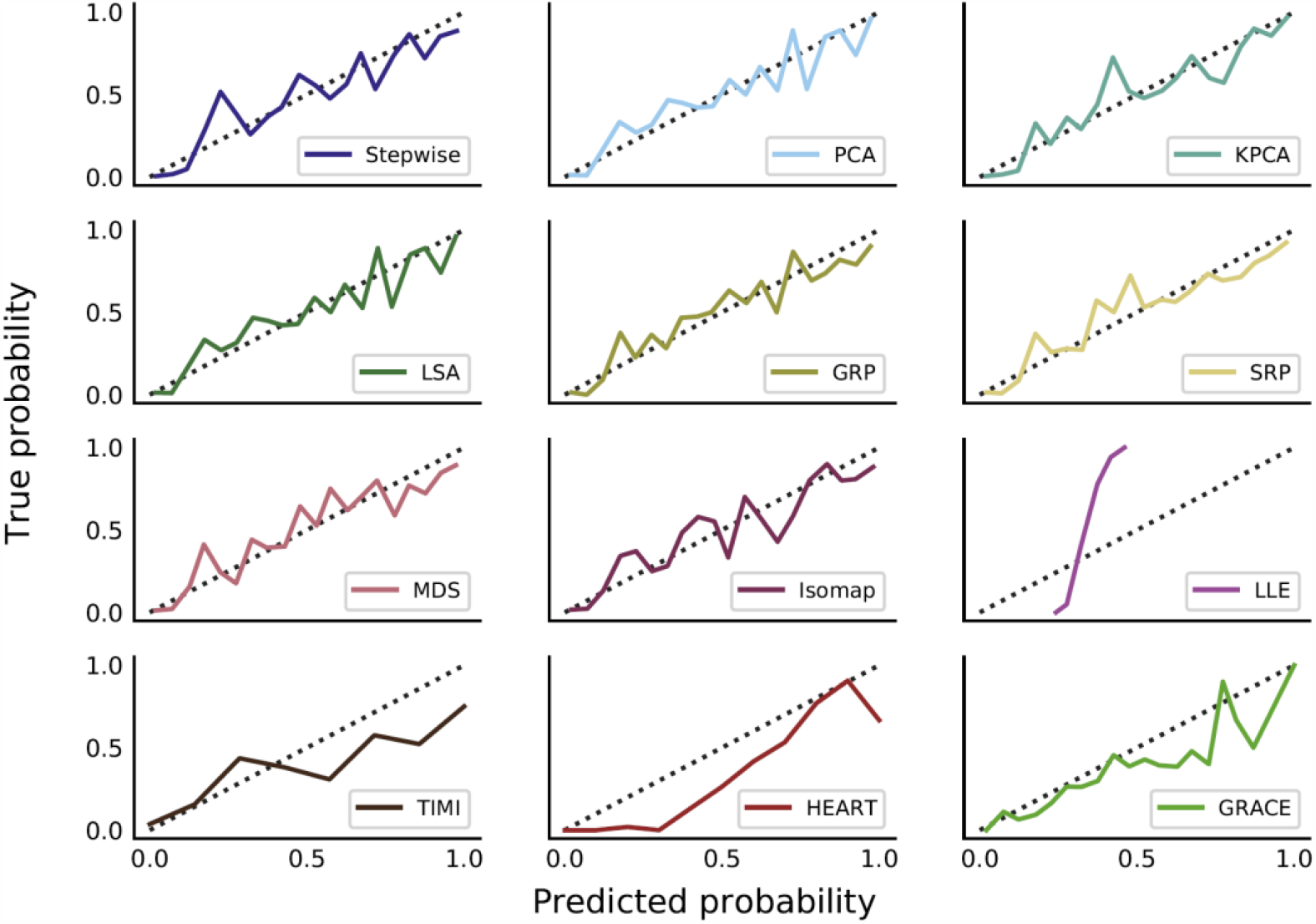
Calibration curves (based on the optimal number of dimensions) generated by the stepwise model, eight dimensionality reduction models, and three clinical scores.

Figure 5 shows ROC curves of HRnV models without using cardiac troponin. At feature dimensions of 13, 21, 13, 29, 24, 17, 18, and 18, the highest AUC values of PCA, KPCA, LSA, GRP, SRP, MDS, Isomap, and LLE were 0.852, 0.852, 0.852, 0.852, 0.851, 0.852, 0.845, and 0.849, respectively. The stepwise model without troponin yielded an AUC of 0.834 compared to 0.887 with troponin. All HRnV-based prediction models outperformed both the TIMI and GRACE scores, while achieving marginally better results than the HEART score.

**Figure 5:**
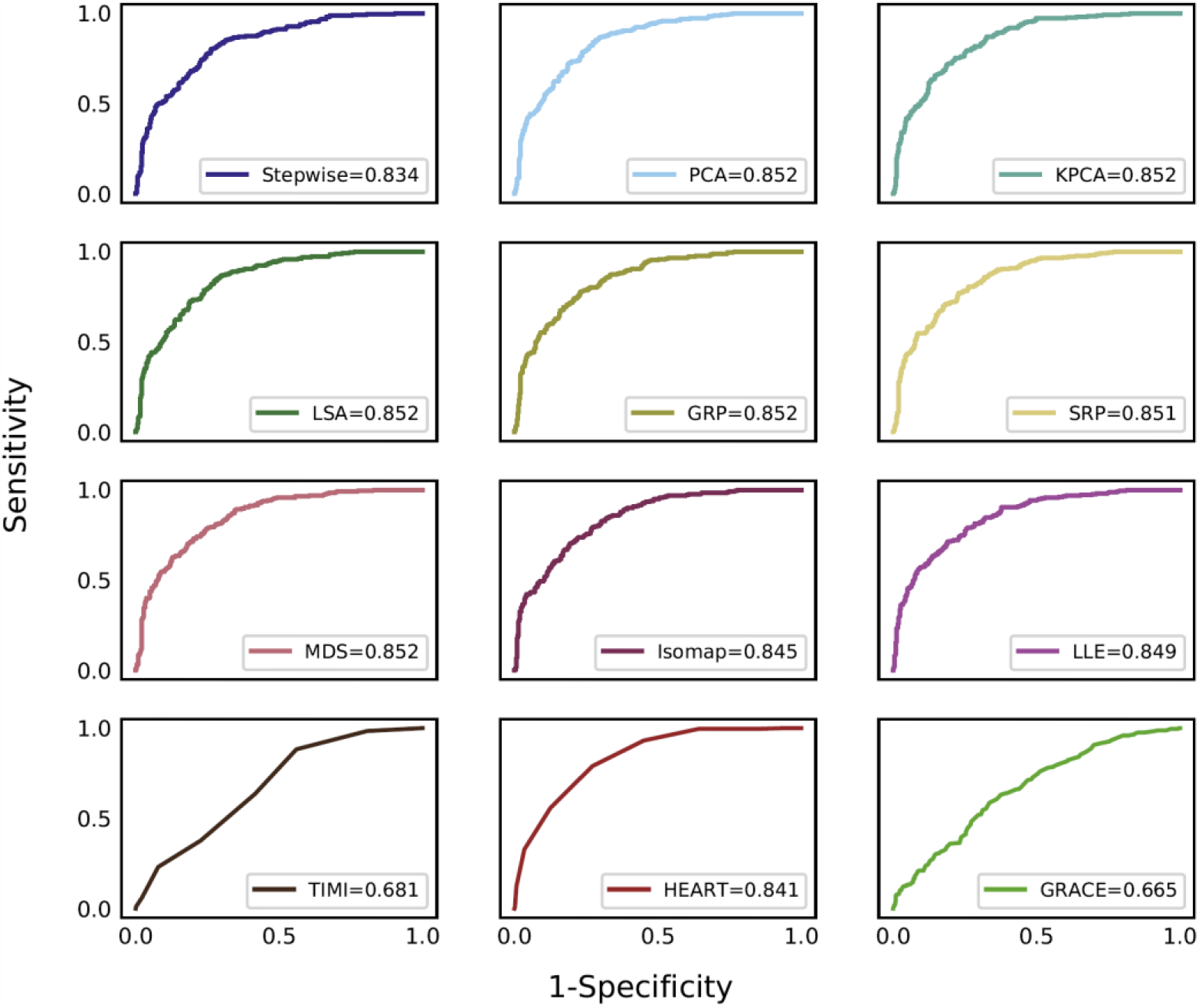
ROC curves (based on the optimal number of dimensions) generated by the stepwise model, eight dimensionality reduction models, and three clinical scores, where the heart rate n-variability based prediction models were built without using cardiac troponin.

## Discussion

In this study, we showed that machine learning dimensionality reduction yielded only marginal, non-significant improvements compared to stepwise variable selection in predicting the risk of 30-day MACE among chest pain patients in the ED. This corroborates with similar observations that traditional statistical methods can perform comparably to machine learning algorithms [52, 53]. Among the models integrated with troponin, the MDS model had the best performance (AUC of 0.901, 95% CI 0.874-0.928), which slightly outperformed the stepwise model (AUC of 0.887, 95% CI 0.859-0.916). Among the models that did not include troponin, PCA, KPCA, LSA, GRP, and MDS performed equally well, achieving an AUC of 0.852, compared with the stepwise model without troponin which had an AUC of 0.834. In general, traditional stepwise approach was proved to be equally well as machine learning dimensionality reduction methods in risk prediction, while benefiting from model simplicity, transparency, and interpretability that are desired in real-world clinical practice.

High-dimensional data suffers from the curse of dimensionality, which refers to the exponentially increasing sparsity of data and sample size required to estimate a function to a given accuracy as dimensionality increases [54]. Dimensionality reduction has successfully mitigated the curse of dimensionality in the analysis of high-dimensional data in various domains such as computational biology and bioinformatics [31, 32]. However, clinical predictive modelling typically considers relatively few features, limiting the effects of the curse of dimensionality. This may account for the relatively limited benefit of dimensionality reduction in our analysis.

Additionally, despite marginal improvements in performance compared to stepwise variable selection, transparency and interpretability of machine learning dimensionality reduction-based methods are limited as they rely on complex algorithmic transformations of variables to a lower-dimensional space, leading to obstacles in the adoption of such models in real-world clinical settings. In contrast, traditional biostatistical approaches like logistic regression with stepwise variable selection deliver a simple and transparent model, in which the absolute and relative importance of each variable can be easily interpreted and explained from the odds ratio. Marginal improvements in performance should be weighed against these limitations in interpretability, which is an important consideration in clinical predictive modelling.

Comparing the eight dimensionality reduction algorithms, PCA and LSA use common linear algebra techniques to learn to create principal components in a compressed data space, while MDS, Isomap, and LLE are nonlinear, manifold learning-based dimensionality reduction methods. As observed from our results, complex nonlinear algorithms did not show obvious advantage over simple PCA and LSA methods in enhancing the predictive performance. Yet, nonlinear algorithms are more computationally complex and require more computing memory. For example, KPCA and Isomap have computational complexity of *O*(*n*^3^) and memory complexity of *O*(*n*^2^), while PCA has computational complexity of 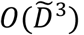 and memory complexity of 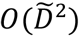 [39]. In applications of clinical predictive modelling, *n* — the number of patients — is usually larger than 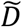— the number of variables; in our study, *n* is 795 and 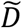 is 29 or 30, depending on the inclusion of troponin. This suggests that linear algorithms may be preferred due to reduced computational complexity and memory while retaining comparable performance. Another observation in this study was that the impact of preselection (as shown in Figure 1) on predictive performance was more substantial than that of dimensionality reduction, indicating the importance of choosing statistically significant candidate variables.

Our study also reiterates the value of HRnV-based prediction models for chest pain risk stratification. Among chest pain risk stratification tools in the ED, clinical scores like HEART, TIMI, and GRACE are currently the most widely adopted and validated [55, 56]. However, a common barrier to quick risk prediction using these traditional clinical scores is the requirement of cardiac troponin, which can take hours to obtain. To address these difficulties, machine learning-based predictive models that integrate HRV measures and clinical parameters have been proposed [17, 22, 25, 26], including our development of HRnV, a novel alternative measure to HRV that has shown promising results in predicting 30-day MACE [28], which was the stepwise model in this paper. Both the dimensionality reduction-based predictive models and the stepwise model with troponin presented superior performance than HEART, TIMI, and GRACE scores. When troponin was not used, several dimensionality reduction-based models such as PCA, KPCA, and MDS still yielded marginally better performance than the original HEART score, while benefiting from generating the predictive scores in merely five to six minutes.

Additionally, Table 3 shows that all HRnV-based predictive models had higher specificities than the HEART score while all HRnV-based models except Isomap also improved on the already high sensitivity of the HEART score [21, 57]. The specificities of KPCA, Isomap, and MDS were significantly higher by an absolute value of almost 10 percent. Substantial improvements to the specificity of MACE predictive models may reduce unnecessary admission and thus minimise costs and resource usage [5]. This is particularly relevant in low-resource settings, for example, the overburdened EDs in the current coronavirus disease 2019 (COVID-19) pandemic, where novel methods in resource allocation and risk stratification could alleviate the strain on healthcare resources[58].

There remains a need for further investigation into methods that utilise information from the full set of HRV and HRnV variables. From 174 variables in the initial data set, dimensionality reduction performed the best with a preselection of 30 variables, of which 19 were HRV and HRnV parameters. That is, majority of the newly constructed HRnV parameters were removed based on the strict significance threshold of p<0.02 on univariable analysis. Therefore, novel HRnV measures were not fully used in prediction models of 30-day MACE, leaving room for further investigation of alternative ways of using them. Moving forward, it may be valuable to develop and evaluate deep learning frameworks [59] to synthesize novel low-dimensional representations of multidimensional information.

We acknowledge the following limitations of this study. First, the clinical application (i.e. risk stratification of ED chest pain patients) was only one example of clinical predictive modelling, thus our conclusion on the effectiveness of machine learning dimensionality reduction algorithms may not be generalizable to other applications, particularly those with a larger number of variables. Secondly, only eight most used dimensionality reduction algorithms were investigated, while many other methods are available. Last, we did not build a workable predictive model for risk stratification of ED chest pain patients, although the HRnV-based models showed promising results compared with existing clinical scores. We aim to further investigate deep representations of the input variables (both HRnV parameters and clinical information) to achieve more rapid prediction and significantly improve predictive performance.

## Conclusions

To our knowledge, this was the first study to compare machine learning dimensionality reduction with conventional stepwise variable selection for risk stratification of chest pain patients in the ED. We found that HRnV-based models using stepwise logistic regression performed better than existing chest pain scores for predicting MACE, with only marginal improvements using machine learning dimensionality reduction. This demonstrated the clinical applicability of the traditional stepwise prediction model due to its simplicity, transparency, and interpretability. To fully utilise the information (i.e. variables) in building high-performing predictive models, we suggest that additional investigations into methods to better represent the large amount of HRnV variables should be conducted. Furthermore, explainable artificial intelligence deserves in-depth exploration such that the derived models are interpretable by clinicians.

## Data Availability

The datasets used and/or analyzed during the current study are available from the corresponding author on reasonable request.

## List of abbreviations

ACS: acute coronary syndrome
AMI: acute myocardial infarction
AUC: area under the curve
ApEn: approximate entropy
CI: confidence intervals
CABG: coronary artery bypass graft
COVID-19: coronavirus disease 2019
DFA: detrended fluctuation analysis
ECG: electrocardiogram
EHR: electronic health records
ED: emergency department
GRP: Gaussian random projection
GRACE: global registry of acute coronary events
HRnV: heart rate n-variability
HRV: heart rate variability
HF: high frequency
HEART: History, ECG, Age, Risk factors, and initial Troponin
IQR: interquartile range
KPCA: kernel principal component analysis
LSA: latent semantic analysis
LLE: locally linear embedding
LF: low frequency
MACE: major adverse cardiac events
Mean NN: average of R-R intervals
MDS: multidimensional scaling
NPV: negative predictive value
NN50: the number of times that the absolute difference between 2 successive R-R intervals exceeds 50 ms
NN50*n*: the number of times that the absolute difference between 2 successive RR_*n*_I/RR_*n*_I_*m*_ sequences exceeds 50×*n* ms
PACS: patient acuity category scale PCI, percutaneous coronary intervention
pNN50: NN50 divided by the total number of R-R intervals
pNN50*n*: NN50*n* divided by the total number of RR_*n*_I/RR_*n*_I_*m*_ sequences PPV, positive predictive value
PCA: principal component analysis
ROC: receiver operating characteristic
RMSSD: square root of the mean squared differences between R-R intervals
RRI: R-R interval
SampEn: sample entropy
SD: standard deviation
SDNN: standard deviation of R-R intervals
SRP: sparse random projection
STEMI: ST-elevation myocardial infarction
TIMI: thrombolysis in myocardial infarction
VLF: very low frequency

## Declarations

### Ethics approval and consent to participate

The ethical approval was obtained from the Centralized Institutional Review Board (CIRB, Ref: 2014/584/C) of SingHealth, in which patient consent was waived.

### Consent for publication

Not applicable.

### Competing interests

NL and MEHO hold patents related to using heart rate variability and artificial intelligence for medical monitoring. NL, ZXK, DG, and MEHO are currently advisers to TIIM SG. The other authors report no conflicts.

### Funding

This work was supported by the Duke-NUS Signature Research Programme funded by the Ministry of Health, Singapore. The funder of the study had no role in study design, data collection, data analysis, data interpretation, or writing of the report.

### Authors’ contributions

NL conceived the study and supervised the project. NL and MLC performed the analyses and drafted the manuscript. NL, MLC, ZXK, SLL, AFWH, DG, and MEHO made substantial contributions to results interpretation and critical revision of the manuscript. All authors read and approved the final manuscript.

## Acknowledgements

We would like to thank and acknowledge the contributions of doctors, nurses, and clinical research coordinators from the Department of Emergency Medicine, Singapore General Hospital.

## References

1. Long B, Koyfman A: Best Clinical Practice: Current Controversies in Evaluation of Low-Risk Chest Pain-Part 1. J Emerg Med 2016, 51(6):668-676.

2. Long B, Koyfman A: Best Clinical Practice: Current Controversies in the Evaluation of Low-Risk Chest Pain with Risk Stratification Aids. Part 2. J Emerg Med 2017, 52(1):43–51.

3. Januzzi JL, Jr., McCarthy CP: Evaluating Chest Pain in the Emergency Department: Searching for the Optimal Gatekeeper. Journal of the American College of Cardiology 2018, 71(6):617–619.

4. Pope JH, Aufderheide TP, Ruthazer R, Woolard RH, Feldman JA, Beshansky JR, Griffith JL, Selker HP: Missed diagnoses of acute cardiac ischemia in the emergency department. N Engl J Med 2000, 342(16):1163–1170.

5. Hollander JE, Than M, Mueller C: State-of-the-Art Evaluation of Emergency Department Patients Presenting With Potential Acute Coronary Syndromes. Circulation 2016, 134(7):547–564.

6. Antman EM, Cohen M, Bernink PJ, McCabe CH, Horacek T, Papuchis G, Mautner B, Corbalan R, Radley D, Braunwald E: The TIMI risk score for unstable angina/non-ST elevation MI: A method for prognostication and therapeutic decision making. JAMA 2000, 284(7):835–842.

7. Morrow DA, Antman EM, Charlesworth A, Cairns R, Murphy SA, de Lemos JA, Giugliano RP, McCabe CH, Braunwald E: TIMI risk score for ST-elevation myocardial infarction: A convenient, bedside, clinical score for risk assessment at presentation: An intravenous nPA for treatment of infarcting myocardium early II trial substudy. Circulation 2000, 102(17):2031–2037.

8. Fox KA, Dabbous OH, Goldberg RJ, Pieper KS, Eagle KA, Van de Werf F, Avezum A, Goodman SG, Flather MD, Anderson FA, Jr. et al: Prediction of risk of death and myocardial infarction in the six months after presentation with acute coronary syndrome: prospective multinational observational study (GRACE). BMJ (Clinical research ed) 2006, 333(7578):1091.

9. Six AJ, Backus BE, Kelder JC: Chest pain in the emergency room: value of the HEART score. Netherlands heart journal : monthly journal of the Netherlands Society of Cardiology and the Netherlands Heart Foundation 2008, 16(6):191–196.

10. Backus BE, Six AJ, Kelder JC, Bosschaert MA, Mast EG, Mosterd A, Veldkamp RF, Wardeh AJ, Tio R, Braam R et al: A prospective validation of the HEART score for chest pain patients at the emergency department. International Journal of Cardiology 2013, 168(3):2153–2158.

11. Six AJ, Cullen L, Backus BE, Greenslade J, Parsonage W, Aldous S, Doevendans PA, Than M: The HEART score for the assessment of patients with chest pain in the emergency department: a multinational validation study. Critical pathways in cardiology 2013, 12(3):121–126.

12. Chen X-H, Jiang H-L, Li Y-M, Chan CPY, Mo J-R, Tian C-W, Lin P-Y, Graham CA, Rainer TH: Prognostic values of 4 risk scores in Chinese patients with chest pain: Prospective 2-centre cohort study. Medicine 2016, 95(52):e4778.

13. Jain T, Nowak R, Hudson M, Frisoli T, Jacobsen G, McCord J: Short- and Long-Term Prognostic Utility of the HEART Score in Patients Evaluated in the Emergency Department for Possible Acute Coronary Syndrome. Critical pathways in cardiology 2016, 15(2):40–45.

14. Sakamoto JT, Liu N, Koh ZX, Fung NX, Heldeweg ML, Ng JC, Ong ME: Comparing HEART, TIMI, and GRACE scores for prediction of 30-day major adverse cardiac events in high acuity chest pain patients in the emergency department. International Journal of Cardiology 2016, 221:759–764.

15. Sun BC, Laurie A, Fu R, Ferencik M, Shapiro M, Lindsell CJ, Diercks D, Hoekstra JW, Hollander JE, Kirk JD et al: Comparison of the HEART and TIMI Risk Scores for Suspected Acute Coronary Syndrome in the Emergency Department. Critical pathways in cardiology 2016, 15(1):1–5.

16. Poldervaart JM, Langedijk M, Backus BE, Dekker IMC, Six AJ, Doevendans PA, Hoes AW, Reitsma JB: Comparison of the GRACE, HEART and TIMI score to predict major adverse cardiac events in chest pain patients at the emergency department. International Journal of Cardiology 2017, 227:656–661.

17. Sakamoto JT, Liu N, Koh ZX, Guo D, Heldeweg MLA, Ng JCJ, Ong MEH: Integrating heart rate variability, vital signs, electrocardiogram, and troponin to triage chest pain patients in the ED. Am J Emerg Med 2017.

18. Engel J, Heeren MJ, van der Wulp I, de Bruijne MC, Wagner C: Understanding factors that influence the use of risk scoring instruments in the management of patients with unstable angina or non-ST-elevation myocardial infarction in the Netherlands: a qualitative study of health care practitioners’ perceptions. BMC health services research 2014, 14:418.

19. Wu WK, Yiadom MY, Collins SP, Self WH, Monahan K: Documentation of HEART score discordance between emergency physician and cardiologist evaluations of ED patients with chest pain. Am J Emerg Med 2017, 35(1):132–135.

20. Ras M, Reitsma JB, Hoes AW, Six AJ, Poldervaart JM: Secondary analysis of frequency, circumstances and consequences of calculation errors of the HEART (history, ECG, age, risk factors and troponin) score at the emergency departments of nine hospitals in the Netherlands. BMJ Open 2017, 7(10):e017259.

21. Laureano-Phillips J, Robinson RD, Aryal S, Blair S, Wilson D, Boyd K, Schrader CD, Zenarosa NR, Wang H: HEART Score Risk Stratification of Low-Risk Chest Pain Patients in the Emergency Department: A Systematic Review and Meta-Analysis. Ann Emerg Med 2019, 74(2):187–203.

22. Ong MEH, Goh K, Fook-Chong S, Haaland B, Wai KL, Koh ZX, Shahidah N, Lin Z: Heart rate variability risk score for prediction of acute cardiac complications in ED patients with chest pain. Am J Emerg Med 2013, 31(8):1201–1207.

23. Rajendra Acharya U, Paul Joseph K, Kannathal N, Lim CM, Suri JS: Heart rate variability: a review. Med Biol Eng Comput 2006, 44(12):1031–1051.

24. Liu N, Koh ZX, Chua ECP, Tan LML, Lin Z, Mirza B, Ong MEH: Risk scoring for prediction of acute cardiac complications from imbalanced clinical data. IEEE J Biomed Health Inform 2014, 18(6):1894–1902.

25. Liu N, Lin Z, Cao J, Koh ZX, Zhang T, Huang G-B, Ser W, Ong MEH: An intelligent scoring system and its application to cardiac arrest prediction. IEEE Transactions on Information Technology in Biomedicine 2012, 16(6):1324–1331.

26. Heldeweg ML, Liu N, Koh ZX, Fook-Chong S, Lye WK, Harms M, Ong ME: A novel cardiovascular risk stratification model incorporating ECG and heart rate variability for patients presenting to the emergency department with chest pain. Crit Care 2016, 20(1):179.

27. Liu N, Koh ZX, Goh J, Lin Z, Haaland B, Ting BP, Ong MEH: Prediction of adverse cardiac events in emergency department patients with chest pain using machine learning for variable selection. BMC Med Inform Decis Mak 2014, 14:75.

28. Liu N, Guo D, Koh ZX, Ho AFW, Xie F, Tagami T, Sakamoto JT, Pek PP, Chakraborty B, Lim SH et al: Heart rate n-variability (HRnV) and its application to risk stratification of chest pain patients in the emergency department. BMC Cardiovasc Disord 2020, 20(1):168.

29. Meloun M, Militký J, Hill M, Brereton RG: Crucial problems in regression modelling and their solutions. Analyst 2002, 127(4):433–450.

30. Dormann CF, Elith J, Bacher S, Buchmann C, Carl G, Carré G, Marquéz JRG, Gruber B, Lafourcade B, Leitão PJ et al: Collinearity: a review of methods to deal with it and a simulation study evaluating their performance. Ecography 2013, 36(1):27–46.

31. Gui J, Andrew AS, Andrews P, Nelson HM, Kelsey KT, Karagas MR, Moore JH: A Robust Multifactor Dimensionality Reduction Method for Detecting Gene-Gene Interactions with Application to the Genetic Analysis of Bladder Cancer Susceptibility. Annals of Human Genetics 2011, 75(1):20–28.

32. Ritchie MD, Hahn LW, Roodi N, Bailey LR, Dupont WD, Parl FF, Moore JH: Multifactor-Dimensionality Reduction Reveals High-Order Interactions among Estrogen-Metabolism Genes in Sporadic Breast Cancer. The American Journal of Human Genetics 2001, 69(1):138–147.

33. Akhbardeh A, Jacobs MA: Comparative analysis of nonlinear dimensionality reduction techniques for breast MRI segmentation. Medical Physics 2012, 39(4):2275–2289.

34. Balvay D, Kachenoura N, Espinoza S, Thomassin-Naggara I, Fournier LS, Clement O, Cuenod C-A: Signal-to-Noise Ratio Improvement in Dynamic Contrastenhanced CT and MR Imaging with Automated Principal Component Analysis Filtering. Radiology 2011, 258(2):435–445.

35. Tarvainen MP, Cornforth DJ, Jelinek HF: Principal component analysis of heart rate variability data in assessing cardiac autonomic neuropathy. In: 2014 36th Annual International Conference of the IEEE Engineering in Medicine and Biology Society. 2014: 6667–6670.

36. Vest AN, Da Poian G, Li Q, Liu C, Nemati S, Shah AJ, Clifford GD: An open source benchmarked toolbox for cardiovascular waveform and interval analysis. Physiol Meas 2018, 39(10):105004.

37. Maimon O, Rokach L: Data Mining and Knowledge Discovery Handbook: Springer Publishing Company, Incorporated; 2010.

38. Zhang Z: Variable selection with stepwise and best subset approaches. Ann Transl Med 2016, 4(7):136.

39. van der Maaten LJP, Postma EO, van den Herik HJ: Dimensionality Reduction: A Comparative Review. In: Tilburg University Technical Report TiCC-TR 2009-005. Tilburg University; 2009.

40. Nguyen LH, Holmes S: Ten quick tips for effective dimensionality reduction. PLoS Comput Biol 2019, 15(6):e1006907–e1006907.

41. Liu N, Wang H: Weighted principal component extraction with genetic algorithms. Applied Soft Computing 2012, 12(2):961–974.

42. Pan Y, Ge SS, Al Mamun A: Weighted locally linear embedding for dimension reduction. Pattern Recognition 2009, 42(5):798–811.

43. Artoni F, Delorme A, Makeig S: Applying dimension reduction to EEG data by Principal Component Analysis reduces the quality of its subsequent Independent Component decomposition. Neuroimage 2018, 175:176–187.

44. Diamantaras KI, Kung SY: Principal component neural networks: theory and applications: John Wiley & Sons, Inc.; 1996.

45. Schölkopf B, Smola AJ, Müller K-R: Kernel principal component analysis. In: Advances in kernel methods: support vector learning. MIT Press; 1999: 327–352.

46. Landauer TK, Foltz PW, Laham D: An introduction to latent semantic analysis. Discourse Processes 1998, 25(2-3):259–284.

47. Dasgupta S: Experiments with random projection. In: Proceedings of the Sixteenth conference on Uncertainty in artificial intelligence. Stanford, California: Morgan Kaufmann Publishers Inc.; 2000: 143–151.

48. Li P, Hastie TJ, Church KW: Very sparse random projections. In: Proceedings of the 12th ACM SIGKDD international conference on Knowledge discovery and data mining. Philadelphia, PA, USA: Association for Computing Machinery; 2006: 287–296.

49. Mead A: Review of the Development of Multidimensional Scaling Methods. Journal of the Royal Statistical Society Series D (The Statistician) 1992, 41(1):27–39.

50. Tenenbaum JB, de Silva V, Langford JC: A global geometric framework for nonlinear dimensionality reduction. Science (New York, NY) 2000, 290(5500):2319–2323.

51. Roweis ST, Saul LK: Nonlinear dimensionality reduction by locally linear embedding. Science (New York, NY) 2000, 290(5500):2323–2326.

52. Gravesteijn BY, Nieboer D, Ercole A, Lingsma HF, Nelson D, van Calster B, Steyerberg EW: Machine learning algorithms performed no better than regression models for prognostication in traumatic brain injury. Journal of clinical epidemiology 2020, 122:95–107.

53. Nusinovici S, Tham YC, Chak Yan MY, Wei Ting DS, Li J, Sabanayagam C, Wong TY, Cheng CY: Logistic regression was as good as machine learning for predicting major chronic diseases. Journal of clinical epidemiology 2020, 122:56–69.

54. Lee JA, Verleysen M: Nonlinear dimensionality reduction. New York: Springer; 2007.

55. D’Ascenzo F, Biondi-Zoccai G, Moretti C, Bollati M, Omedè P, Sciuto F, Presutti DG, Modena MG, Gasparini M, Reed MJ et al: TIMI, GRACE and alternative risk scores in Acute Coronary Syndromes: A meta-analysis of 40 derivation studies on 216,552 patients and of 42 validation studies on 31,625 patients. Contemporary Clinical Trials 2012, 33(3):507–514.

56. Liu N, Ng JCJ, Ting CE, Sakamoto JT, Ho AFW, Koh ZX, Pek PP, Lim SH, Ong MEH: Clinical scores for risk stratification of chest pain patients in the emergency department: an updated systematic review. Journal of Emergency and Critical Care Medicine 2018, 2:16–16.

57. Byrne C, Toarta C, Backus B, Holt T: The HEART score in predicting major adverse cardiac events in patients presenting to the emergency department with possible acute coronary syndrome: protocol for a systematic review and meta-analysis. Systematic Reviews 2018, 7(1).

58. Liu N, Chee ML, Niu C, Pek PP, Siddiqui FJ, Ansah JP, Matchar DB, Lam SSW, Abdullah HR, Chan A et al: Coronavirus disease 2019 (COVID-19): an evidence map of medical literature. BMC medical research methodology 2020, 20(1):177.

59. Xie J, Girshick R, Farhadi A: Unsupervised deep embedding for clustering analysis. In: Proceedings of the 33rd International Conference on International Conference on Machine Learning - Volume 48. New York, NY, USA: JMLR.org; 2016: 478–487.

